# Prediction of Stroke Risk in Patients with Atrial Fibrillation: Comorbidities vs Temporal AF Behavior

**DOI:** 10.64898/2025.12.18.25342623

**Authors:** Eli Tsakiris, Mario Mekhael, Yuxuan Gu, Christian Massad, Ghassan Bidaoui, Yishi Jia, Yingshuo Liu, Mohammad Montaser Atasi, Yara Menassa, Michel Abou Khalil, Carlo El Khoury, Maximilian Moersdorf, Chanho Lim, Amitabh C. Pandey, Han Feng, Nassir Marrouche

## Abstract

**Background:** There are controversies about whether atrial fibrillation (AF) type, paroxysmal (PaAF) vs persistent (PeAF), affects stroke risk. The type of AF is still not included in most risk stratification tools.

**Objective:** We aim to assess differences in stroke outcomes between PaAF and PeAF patients in both low- and high-CHA_2_DS_2_-VASc groups.

**Methods:** We conducted an epidemiological study of all patients admitted to Tulane Medical Center with the diagnosis of AF from January 2010 to March 2020. Data were extracted from the regional US electronic medical records database, Research Action for Health Network (REACHnet), for all patients aged 18 years or older with a diagnosis of AF. Patients were divided into four groups: a low CHA_2_DS_2_-VASc score was defined as CHA_2_DS_2_-VASc < 2 in women and CHA_2_DS_2_-VASc < 1 in men. PeAF was defined as a patient with at least one episode of AF lasting 7 days or more. PaAF was defined as a patient with AF with no episode lasting more than 7 days. The outcome of the study was an ischemic stroke event or a transient ischemic attack that occurred after the diagnosis of AF. Kaplan-Meier curves and the log-rank test were used to compare the study outcomes across all four groups. Multivariable Cox regression was performed to adjust for the use of anticoagulants.

**Results:** A total of 1,079 patients were included in the study. 576 patients had PaAF and high CHA_2_DS_2_-VASc, 215 had PaAF and low CHA_2_DS_2_-VASc, 214 had PeAF and high CHA_2_DS_2_-VASc, and 74 patients were PeAF, and low CHA_2_DS_2_-VASc. Patients were followed up over 5 years. Based on the Log-rank test, there were significant differences among the four groups (p < 0.001). After adjusting for anticoagulants, patients with high CHA_2_DS_2_-VASc appeared to have more strokes on follow-up than patients with low CHA_2_DS_2_-VASc, independent of AF type and anticoagulation prescription. For the Cox model, when the PaAF High CHA_2_DS_2_-VASc group was used as the reference, both low CHA_2_DS_2_-VASc groups, PaAF (0.032 [0.012–0.081], p < 0.001) and PeAF (0.032 [0.008–0.135], p < 0.001), had a lower risk of stroke. However, there was no difference in stroke when the reference group was compared to high CHA_2_DS_2_-VASc, PeAF (1.169 [0.866 – 1.576], p=0.308).

**Conclusion:** In our database, the CHA_2_DS_2_-VASc score remained superior to the type of AF when predicting stroke outcome. Type of AF did not affect stroke outcome even after adjusting for CHA_2_DS_2_-VASc and for anticoagulation prescription.

## 1. Introduction

Stroke is the most feared complication of atrial fibrillation (AF).^1^ When a stroke occurs, it can either be deadly or associated with debilitating consequences for patients. ^2^ Stroke in AF can be prevented using oral anticoagulation (OAC). However, this may be associated with bleeding risk, which can also be fatal. CHA_2_DS_2_-VASc is the standard risk stratification score used to determine OAC decisions for patients with AF, as per consensus guidelines.^3,4^ Although it accounts for common comorbidities, it does not consider the type of AF, which may influence thromboembolic risk.^3,4^ Previous literature has reported conflicting results regarding whether the persistent AF (PeAF) type confers a higher risk of stroke compared with paroxysmal AF (PaAF), with no cohesive conclusion. Some large randomized clinical trials have suggested that, in patients on OAC, PeAF is associated with a higher absolute risk; however, the relative effect is small after adjusting for comorbidities.^5,6^ Pooled analysis of the Stroke Prevention in AF (SPAF) trials demonstrated a comparable risk of stroke in paroxysmal and permanent AF. ^7–9^. These findings are also supported by data from the Euro Heart Survey, which also showed no difference in stroke between the two types.^10^ Accordingly, this evidence explains why the type of AF is still not incorporated into the most frequently used clinical risk stratification. In addition, as more data emerge on the quantitative percent of AF burden from wearable devices, a qualitative description, or simply paroxysmal vs. persistent, may prove suboptimal for patient care.

This study aims to examine the independent effect of AF type (persistent or paroxysmal) on ischemic stroke, irrespective of CHA_2_DS_2_-VASc score or OAC prescription.

## 2. Methods

### 2.1 Study Design and Population

This retrospective cohort study included all patients aged ≥18 years admitted to Tulane Medical Center between January 2010 and March 2020 with a diagnosis of AF. Data were extracted from the Research Action for Health Network (REACHnet), a regional electronic medical record database. Diagnoses were reconciled using the appropriate 10th edition of the International Classification of Diseases (ICD) as shown in **Table 1**. Patients were stratified into CHA_2_DS_2_-VASc risk categories based on sex-specific thresholds: low risk (CHA_2_DS_2_-VASc ≤1 in men and ≤2 in women) and high risk (CHA_2_DS_2_-VASc >1 in men and>2 in women). Four groups were thus generated, including each pairing of high/low CHA_2_DS_2_-VASc vs PeAF/PaAF.

**Table 1.**
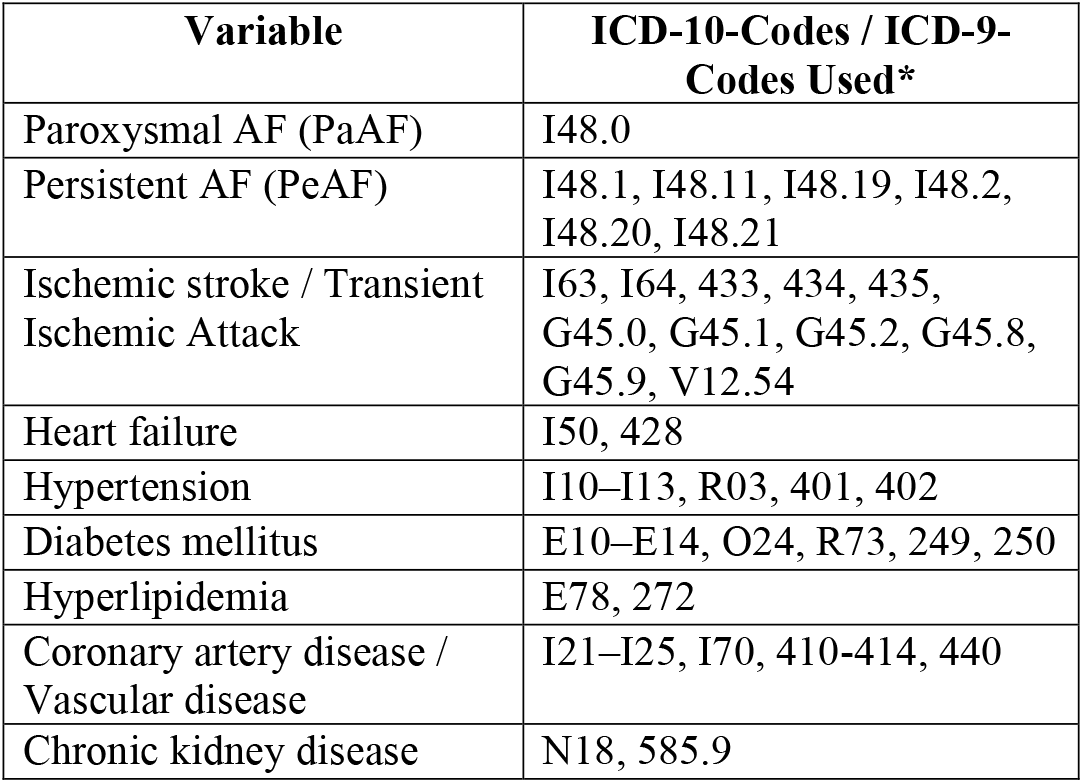
ICD-10-CM Diagnosis Codes Used to Define Exposure, Outcomes, and Covariatess. A description of International Classification of Diseases, 10^th^ Edition (ICD-10) codes used to reconcile various diagnoses used as variables in this study.

| Variable | ICD-10-Codes / ICD-9-Codes Used* |
| --- | --- |
| Paroxysmal AF (PaAF) | I48.0 |
| Persistent AF (PeAF) | I48.1, I48.11, I48.19, I48.2, I48.20, I48.21 |
| Ischemic stroke / Transient Ischemic Attack | I63, I64, 433, 434, 435, G45.0, G45.1, G45.2, G45.8, G45.9, V12.54 |
| Heart failure | I50, 428 |
| Hypertension | I10–I13, R03, 401, 402 |
| Diabetes mellitus | E10–E14, O24, R73, 249, 250 |
| Hyperlipidemia | E78, 272 |
| Coronary artery disease / Vascular disease | I21–I25, I70, 410-414, 440 |
| Chronic kidney disease | N18, 585.9 |

### 2.2 Primary Outcome

The primary outcome was ischemic stroke, defined as either ischemic cerebrovascular accident (CVA) or transient ischemic attack (TIA) occurring after the AF diagnosis.

### 2.3 Statistical Analysis

Baseline demographic and clinical variables were summarized as means and standard deviations for continuous variables, and as counts and percentages for categorical variables. Among the AF type and CHA_2_DS_2_-VASc risk groups, comparisons were conducted using ANOVA and the Wilcoxon test as appropriate for continuous variables, while the Chi-square test was used for categorical variables. Survival analyses were performed using Kaplan–Meier (KM) curves and log-rank tests to compare event-free survival across the four subgroups. Multivariable Cox proportional hazards regression models were used to estimate hazard ratios (HRs) and 95% confidence intervals (CIs) for stroke risk, with adjustment for anticoagulation status. Analyses were performed using R (version 4.5.1)

## 3. Results

A total of 1,079 patients met the inclusion criteria. Of these, 576 (53%) had PaAF and high CHA_2_DS_2_-VASc scores, 215 (20%) had PaAF with low CHA_2_DS_2_-VASc, 214 (20%) had PeAF with high CHA_2_DS_2_-VASc, and 74 (7%) had PeAF with low CHA_2_DS_2_-VASc. The comprehensive baseline demographics for the four groups are presented in **Table 2**. Patients in high CHA_2_DS_2_-VASc groups, as assumed, were older and had a higher burden of comorbidity. Mean ages were 63.75 ± 10.24 for PaAF–high CHA_2_DS_2_-VASc, 58.48 ± 13.72 for PaAF–low CHA_2_DS_2_-VASc, 67.14 ± 8.51 for PeAF–high CHA_2_DS_2_-VASc, and 66.30 ± 10.09 for PeAF– low CHA_2_DS_2_-VASc (*p* < 0.0001). Hypertension prevalence was 92.4%, 50.7%, 92.5%, and 59.5%, respectively (*p* < 0.0001). Diabetes mellitus was present in 68.1%, 4.2%, 68.7%, and 5.4% (*p* < 0.0001). Hyperlipidemia occurred in 71.9%, 34.9%, 72.4%, and 32.4% of the participants (*p* < 0.0001). Female sex distribution was 36.8%, 39.5%, 38.3%, and 21.6% (*p* = 0.041). Notably, only 50.4% of all included patients were White, with variable numbers of African American patients (50.0%, 27.9%, 49.1%, 14.9%) and patients of other races (5.2%, 11.2%, 6.1% and 5.4%, *p* < 0.001) were present in the four groups.

**Table 2.** Baseline Characteristics. Baseline characteristics of 1,079 patients with atrial fibrillation stratified into four groups by AF type (PaAF vs PeAF) and CHA_2_DS_2_-VASc category (low vs high). High-risk groups were older and had substantially greater comorbidity burden, including hypertension, diabetes mellitus, heart failure, chronic kidney disease, and coronary artery disease. Anticoagulation use (warfarin, DOAC, or other agents) varied significantly across groups. Continuous variables are presented as mean ± standard deviation. Categorical variables are presented as sums with percentages in parentheses.

| Characteristic | PaAF, High<br>CHA<sub>2</sub>DS<sub>2</sub>-<br>VASc (n =<br>576) | PaAF, Low<br>CHA<sub>2</sub>DS<sub>2</sub>-<br>VASc<br>(n = 215) | PeAF, High<br>CHA<sub>2</sub>DS<sub>2</sub>-<br>VASc<br>(n = 214) | PeAF, Low<br>CHA<sub>2</sub>DS<sub>2</sub>-<br>VASc<br>(n = 74) | Total<br>(n = 1079) | p value |
| --- | --- | --- | --- | --- | --- | --- |
| Age (SD) | 63.75 (10.24) | 58.48 (13.72) | 67.14 (8.51) | 66.30 (10.09) | 63.55 (11.08) | < 0.0001 |
| Race (%) |  |  |  |  |  | < 0.0001 |
| White | 258 (44.8) | 131 (60.9) | 96 (44.9) | 59 (79.7) | 544 (50.4) |  |
| African<br>American | 288 (50.0) | 60 (27.9) | 105 (49.1) | 11 (14.9) | 464 (43.0) |  |
| Other | 30 (5.2) | 24 (11.2) | 13 (6.1) | 4 (5.4) | 71 (6.6) |  |
| Diabetes<br>mellitus | 392 (68.1) | 9 (4.2) | 147 (68.7) | 4 (5.4) | 552 (51.2) | < 0.0001 |
| Hyperlipidemi<br>a | 414 (71.9) | 75 (34.9) | 155 (72.4) | 24 (32.4) | 668 (61.9) | < 0.0001 |
| Heart failure<br>(CHF) | 350 (60.8) | 17 (7.9) | 151 (70.6) | 8 (10.8) | 526 (48.7) | < 0.0001 |
| Chronic<br>kidney disease | 242 (42.0) | 41 (19.1) | 77 (36.0) | 16 (21.6) | 376 (34.8) | < 0.0001 |
| Coronary<br>artery disease | 230 (39.9) | 4 (1.9) | 81 (37.9) | 0 (0.0) | 315 (29.2) | < 0.0001 |
| CHA<sub>2</sub>DS<sub>2</sub>-<br>VASc Mean<br>(SD) | 3.50 (1.24) | 1.04 (0.71) | 3.65 (1.34) | 0.97 (0.60) | 2.86 (1.59) | < 0.0001 |
| Warfarin | 128 (22.2) | 6 (2.8) | 68 (31.8) | 7 (9.5) | 209 (19.4) | < 0.0001 |
| DOAC | 220 (38.2) | 39 (18.1) | 82 (38.3) | 18 (24.3) | 359 (33.3) | < 0.0001 |
| Other<br>anticoagulant | 184 (31.9) | 90 (41.9) | 35 (16.4) | 13 (17.6) | 322 (29.8) | < 0.0001 |
| Any<br>anticoagulant/<br>antiplatelet | 484 (84.0) | 133 (61.9) | 172 (80.4) | 37 (50.0) | 826 (76.6) | < 0.0001 |

On Kaplan–Meier (KM) analysis, there were significant differences in stroke-free survival between the four subgroups. Patients with high CHA_2_DS_2_-VASc scores, regardless of AF type, experienced higher stroke rates compared with those with low scores. After adjusting for anticoagulation status and baseline comorbidities, the CHA_2_DS_2_-VASc score remained an independent predictor of stroke risk, while AF type did not. KM curves for the four groups are shown in **Figure 1**.

**Figure 1:**
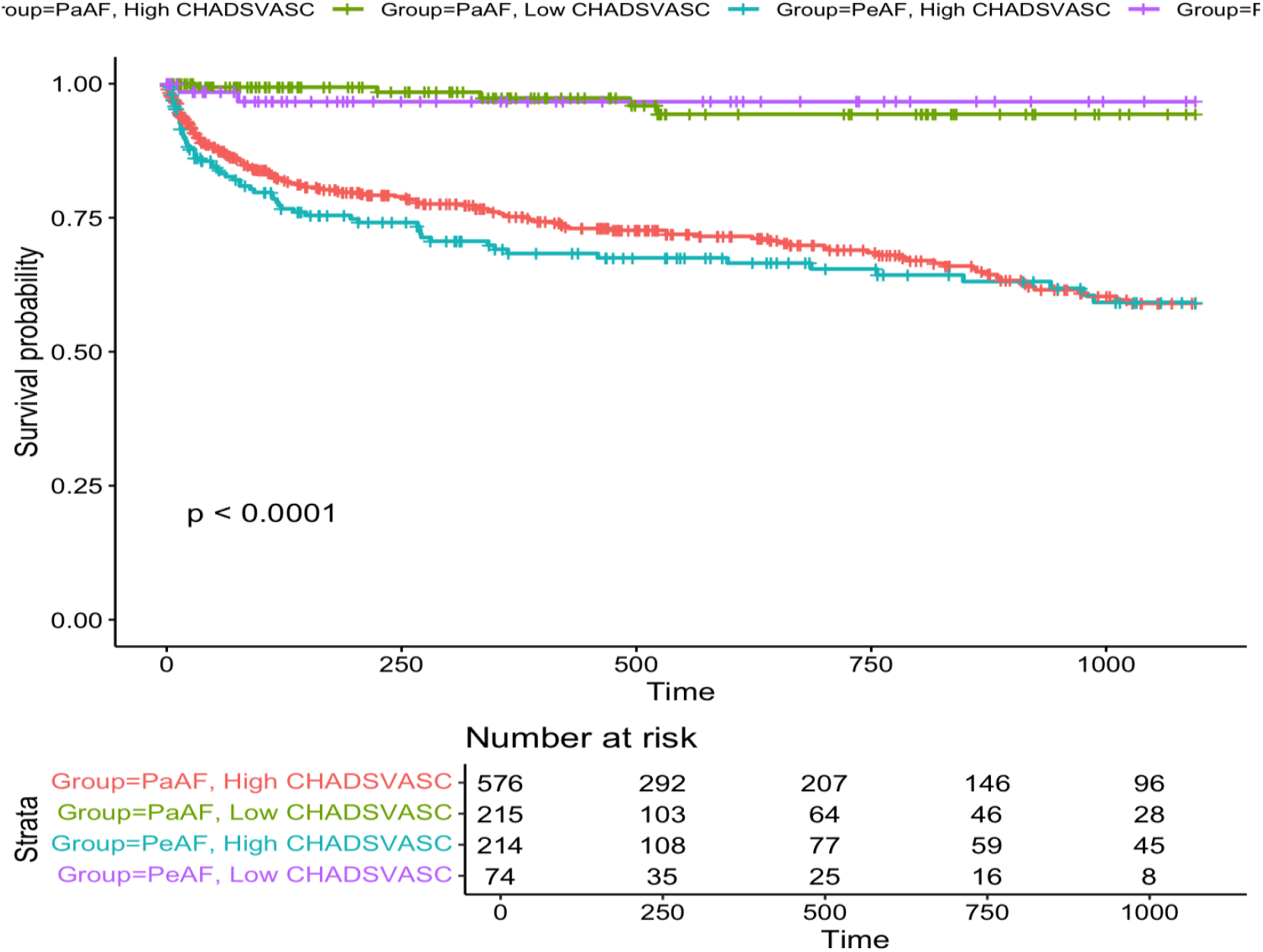
Kaplan–Meier Stroke-Free Survival Stratified by AF Type and CHA_2_DS_2_-VASc Score. This Kaplan–Meier survival curve demonstrates stroke-free survival among four groups stratified by atrial fibrillation (AF) type, paroxysmal (PaAF) or persistent (PeAF), and CHA_2_DS_2_-VASc risk category (low vs high). Patients with high CHA_2_DS_2_-VASc scores exhibited significantly lower stroke-free survival compared with low-risk patients, irrespective of AF type (p < 0.0001). AF type alone did not meaningfully differentiate stroke risk once stratified by CHA_2_DS_2_-VASc category. Number-at-risk counts at pre-specified intervals are shown below the curve.

Using Cox regression and taking the PaAF-high CHA_2_DS_2_-VASc group as reference, the Low CHA_2_DS_2_-VASc groups had significantly lower stroke risk, PaAF (HR 0.032, 95% CI 0.012–0.081; p < 0.001) vs PeAF (HR 0.032, 95% CI 0.008–0.135; p < 0.001). There was no significant difference between PeAF and PaAF in the high CHA_2_DS_2_-VASc group (HR 1.17, 95% CI 0.87–1.58; p = 0.308).

## 4. Discussion

In this study, we note two main findings. First, we found that patients with AF with a high CHA_2_DS_2_-VASc score were more at risk of stroke, independent of the type of AF. Second, those findings persisted despite adjusting for anticoagulation on multivariate analysis.

Importantly, our findings support the concept that stroke risk in atrial fibrillation is driven predominantly by underlying atrial substrate rather than by AF pattern alone. Consistent with this, prior literature suggests that CHA_2_DS_2_-VASc reflects more than just clinical comorbidity burden and functions as a practical surrogate for left atrial myopathy. This relationship is biologically plausible because atrial cardiomyopathy is a multifactorial phenotype shaped by chronic local and systemic inflammation, neurohormonal activation, metabolic dysfunction, and structural remodeling, culminating in atrial fibrosis, impaired mechanical function, and a prothrombotic milieu. Supporting this paradigm, atrial myopathy has been observed to precede and progress independently of incident AF, with evidence of structural and functional abnormalities even among individuals without AF at baseline.^11^ Moreover, systemic inflammatory and metabolic disorders frequently precede AF onset and are thought to both initiate and perpetuate atrial remodeling, providing a mechanistic link between upstream comorbidities and downstream thromboembolic risk.^12^ In this context, it is notable that many of the factors within the CHA_2_DS_2_-VASc score have been independently associated with atrial myopathy, including age, diabetes, hypertension, ischemic heart disease, and heart failure, supporting the score as a reflection of underlying myopathy. ^12–14^ Further, atrial myopathy has been consistently correlated with thromboembolic risk more strongly than AF subtype itself.^15,19^ Direct measures of atrial myopathy (e.g., atrial strain or fibrosis) predict thromboembolic events more accurately than arrhythmia subtype, suggesting that the arrhythmogenic substrate of a diseased atrium contributes to both AF and thromboembolic risk.^20,21^ These findings have been attributed to different consequences of atrial myopathy, including triggering atrial tachyarrhythmia, leading to stiff atrial syndrome and predisposing to left atrial appendage thrombus formation. ^12,14,22^ All in all, the presence of atrial myopathy alters atrial structure, leading to hypercoagulability and increasing stasis, completing Virchow’s triad.^23^ Accordingly, considering the atrial substrate as the primary driver of stroke risk explains the relatively negligible effect of AF subtype compared to CHA_2_DS_2_-VASc.

Despite this, a burden-based effect remains biologically plausible: longer time in AF could amplify stasis and facilitate thrombus formation. This has motivated studies testing whether AF type or burden meaningfully refines risk prediction beyond CHA_2_DS_2_-VASc.

Accordingly, there is interest in whether the inclusion of AF type or burden could improve the accuracy of risk stratification tools, such as CHA_2_DS_2_-VASc. The ARISTOTLE trial, which included 18,201 patients with AF (15.3% PaAF and 84.7% PeAF), examined the efficacy of apixaban and found that the rate of thromboembolic events was higher in the PeAF population, even after robust adjustment.^5^ Subanalysis from the ROCKET-AF trial examining efficacy of rivaroxaban, found similar results with a modestly higher hazard of thromboembolic events in the PeAF population, though notably the study population was very high risk with an average CHA_2_DS_2_-VASc of 5.^6^. The effect of AF type on thromboembolism was mixed in other sub-analyses from seminal OAC trials, with a small effect seen in ENGAGE TIMI AF 38 but no significant effect observed in RE-LY or SPORTIF. ^24–26^ Notably, all patients in each of these trials were on OAC by design, and after adjustment for comorbidities, the independent risks of AF type were markedly attenuated.

Beyond confounding, AF subtype itself is an imprecise exposure metric, given the marked heterogeneity within each category. For example, a patient with frequent, recurrent episodes that all self-terminate within seven days is classified as having PaAF, whereas another patient with a single episode lasting more than seven days is labeled PeAF. However, the former patient may have a higher overall AF burden despite being categorized as paroxysmal (Figure 2), underscoring the need for quantitative measurements to better define disease burden. Growing evidence suggests that AF burden outperforms binary subtype for stroke prediction and may represent a meaningful therapeutic target. ^27,28^ The expansion of wearable and continuous ECG technologies has made longitudinal burden measurement increasingly feasible. ^29–31^ As these tools become ubiquitous, shifting from qualitative AF subtype to quantitative burden may better align classification with clinically relevant outcomes. In parallel, our multivariable analysis supports CHA_2_DS_2_-VASc as a robust predictor of stroke risk regardless of AF type, and extends prior work by evaluating a racially diverse cohort (50.4% White). Continued real-world monitoring will clarify the incremental value of AF burden over subtype for risk stratification.

**Figure 2:**
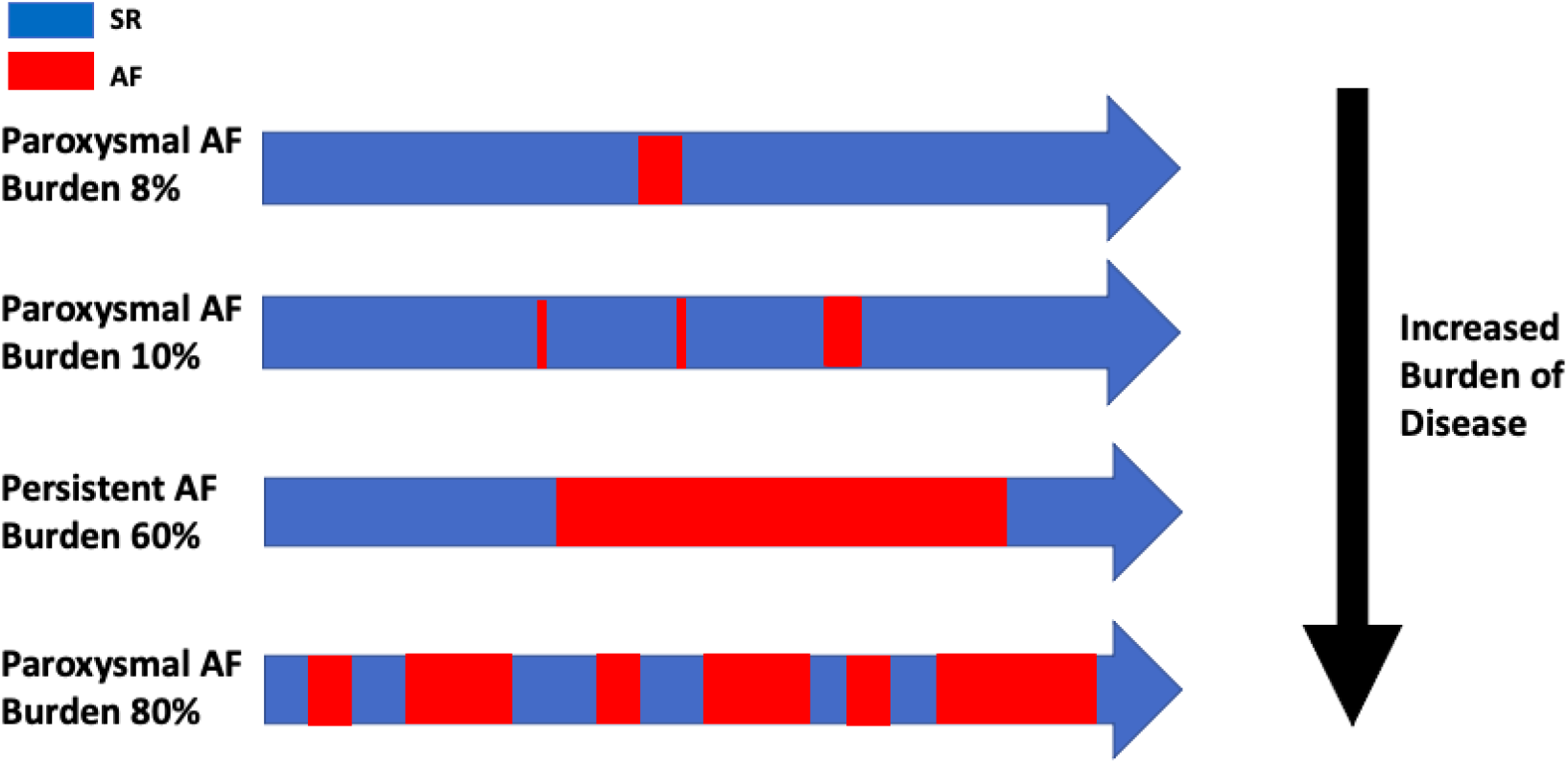
Conceptual Illustration of AF Burden Across Paroxysmal and Persistent AF. Conceptual visualization illustrating the limitations of binary AF type classification (paroxysmal vs persistent) in representing true AF burden. Patients labeled as paroxysmal AF may exhibit widely variable AF burden, in some cases exceeding that of individuals labeled with persistent AF. Increasing AF burden (percentage of time in AF) is shown from top to bottom, emphasizing that qualitative AF type may not reliably reflect total arrhythmia exposure.

### 4.1 Strengths and Limitations

This study provides data from a large, real-world cohort over ten years. The inclusion of patients with and without OAC offers a valuable assessment of stroke risk across different treatment options. The broad range of comorbidity burden, as represented by CHA_2_DS_2_-VASc scores, helps generalize findings more effectively than heavily controlled randomized controlled trials. This retrospective analysis is subject to inherent limitations due to its retrospective nature, reliance on medical record coding, and prescription documentation for OAC, rather than direct confirmation of medication adherence.

## 5. Conclusion

Patients with high CHA_2_DS_2_-VASc score were at a higher risk of stroke, independent of the type of AF. These results reinforce the conclusion that stroke prevention in AF should remain focused on optimizing comorbidities and guideline-directed anticoagulation, rather than on AF subtype classification. Future studies should move beyond qualitative type classifications and instead focus on quantitative, continuous measures of AF burden and their association with clinical outcomes.

## Data Availability

The dataset has been downloaded from here: https://openneuro.org/datasets/ds000220/versions/00002

## Acknowledgments

We acknowledge REACHnet, a PCORnet® Clinical Research Network, for providing the electronic health record data used in this study. The interpretation and conclusions are solely those of the authors.

